## Supplementary Material for "Accuracy of emergency medical service telephone triage of need for an ambulance response in suspected COVID-19: An observational cohort study"

Supplementary Material 1: Summary of AMPDS pandemic Card 36 (full details available: <https://cdn.emergencydispatch.org/iaed/pdf/resource-library/public%20protocols/pub-protocol%2036/NAE%20CC%2036.pdf>)

| **Questions** | **Outcome** |
| --- | --- |
| 1) What is the most prominent complaint? | **Delta Category (immediate response)**  **Category 1/2 UK Ambulance service response**  1) INEFFECTIVE BREATHING with flu-like symptoms 2) DIFFICULTY SPEAKING BETWEEN BREATHS with flu-like symptoms 3) Not alert with flu-like symptoms 4) CHANGING COLOR with flu-like symptoms |
| If breathing a)Do they have difficulty speaking between breathes?  b) Describe their breathing  c)Did they have any flu symptoms prior to this? |  |
| If Chest pain a) Are they 35 and over?  b) Have they had a heart attack or angina previously |  |
| 2) Are they completely alert? |  |
| 3) Are they changing colour? |  |
| 4) Are they having chills or sweats? | **Charlie Category (Ambulance when able)**  **Category 3/4 UK Ambulance service response**  1) Abnormal breathing with single flu-like symptom or Asthma/COPD  2) Abnormal breathing with multiple flu-like symptoms  3) Chest pain/discomfort ≥35 with single flu-like symptom  4) Chest pain/discomfort ≥35 with multiple flu-like symptoms  5) High Risk Factor |
| 5) Are they vomiting? |  |
| 6)Do they have a new cough that recently started? |  |
| 7)Do they have a sore throat? |  |
| 8)Do they have unusual total body aches? |  |
| 9)Do they have a fever? | **Alpha Category (No ambulance dispatched)**  1) Chest pain/discomfort < 35 with single flu-like symptom  2) Chest pain/discomfort < 35 with multiple flu-like symptoms  3) Flu-like symptoms only |
| 10)Do they have a runny or snotty nose? |  |
| 11)Do they have diarrhoea? |  |
| 12)Do they have a headache? |  |
| 13)Do they have any high-risk conditions?  • ≤ 5 years old (not COVID-19) • ≥ 65 years old • Blood disorders • Diabetes • Kidney and liver diseases/disorders • Neurological diseases • Pregnancy (up to 2 weeks after delivery) • Sickle cell disease (sickle cell anaemia) • Weakened immune system |  |

Supplementary Material 2: Multi-variable model predicting false negatives

| **Population Characteristic** | **Level** | **Odds ratio (95% CI)**  **N= 1215** |
| --- | --- | --- |
| **Age (Years)** | 1-year increase | 0.95 (0.94 to 0.97) |
| **Sex** | Female | 1.89 (1.09 to 3.26) |
| **Comorbidity** | Cardiovascular Disease | 0.73 (0.21 to 2.56) |
|  | Chronic Resp. Disease | 0.50 (0.23 to 1.09) |
|  | Diabetes | 1.19 (0.60 to 2.38) |
|  | Hypertension | 0.75 (0.39 to 1.46) |
|  | Immunosuppression  (including steroid use) | 0.73 (0.35 to 1.53) |
|  | Active Malignancy | 0.12 (0.02 to 0.92) |
|  | Obesity | Not included |
|  | Renal Impairment | 2.22 (0.71 to 6.96) |
|  | Smoker | 0.96 (0.52 to 1.80) |
|  | Stroke | 1.93 (0.42 to 8.80) |
| **Number of Drugs Used** | 0 | Reference |
|  | 1-5 | 0.98 (0.45 to 2.15) |
|  | 6-10 | 1.56 (0.55 to 4.44) |
|  | 11 or more | 0.76 (0.08 to 7.42) |
| **Deprivation Index** | 1-2 | Reference |
|  | 3-4 | 0.98 (0.47 to 2.08) |
|  | 5-6 | 0.54 (0.21 to 1.36) |
|  | 7-8 | 1.37 (0.64 to 2.97) |
|  | 9-10 | 0.99 (0.38 to 2.59) |

Supplementary Material 3: Multi-variable model predicting false positives

| **Population Characteristic** | **Level** | **Odds ratio (95% CI)**  **N= 5, 615** |
| --- | --- | --- |
| **Age (Years)** | 1-year increase | 1.05 (1.04 to 1.05) |
| **Sex** | Female | 1.31 (1.15 to 1.49) |
| **Comorbidity** | Cardiovascular Disease | 1.38 (0.85 to 2.24) |
|  | Chronic Resp. Disease | 1.35 (1.13 to 1.60) |
|  | Diabetes | 0.85 (0.67 to 1.09) |
|  | Hypertension | 1.03 (0.84 to 1.26) |
|  | Immunosuppression  (including steroid use) | 1.02 (0.82 to 1.26) |
|  | Active Malignancy | 1.25 (0.72 to 2.18) |
|  | Renal Impairment | 0.80 (0.44 to 1.44) |
|  | Smoker | 0.86 (0.75 to 0.99) |
|  | Stroke | 1.17 (0.59 to 2.31) |
| **Number of Drugs Used** | 0 | Reference |
|  | 1-5 | 0.99 (0.84 to 1.16) |
|  | 6-10 | 1.22 (0.90 to 1.66) |
|  | 11 or more | 1.13 (0.62 to 2.06) |
| **Deprivation Index** | 1-2 | Reference |
|  | 3-4 | 1.21 (1.02 to 1.44) |
|  | 5-6 | 1.23 (1.01 to 1.48) |
|  | 7-8 | 1.29 (1.05 to 1.59) |
|  | 9-10 | 1.32 (1.04 to 1.69) |
