## Supplementary material for "Accuracy of emergency medical service telephone triage of need for an ambulance response in suspected COVID-19: An observational cohort study": Figures

Figure 1: STROBE flow diagram of study population selection

“999” calls received by YAS between 2^nd^ April and 29^th^ June 2020 assessed using COVID-19 pandemic Card 36

*N = 17,905*

Calls for which it was not possible to trace patient’s identity

*N* = *3,225*

NHS Digital routine data sources: APC, CC, ECDS, DEMO, DR & GDPPR

Linkable calls

*N = 14,680*

Multiple calls single patient or excluded patients

*N* = *1,569*

Cohort of individual patients

*N* = *13,111*

Aged<16

*N* = *458*

Final study population

*N* = 12,653
